# Does greater longevity come with lower life disparity in India? A comparative study between Kerala and Delhi

**DOI:** 10.1101/2022.04.26.22274327

**Authors:** Saddaf Naaz Akhtar, Jon Anson

## Abstract

**Background:** Life expectancy and life disparity are two important measures for determining society’s health condition. Over last decades, Indian life expectancy has increased, reaching 69.4 years in 2018, with highest life expectancies being registered in Kerala and Delhi, with 75.3 years for both-sexes. Delhi has overtaken Kerala and is now top-ranking state in life expectancy. However, whether Delhi also has a lower disparity than Kerala is still unknown.

**Objectives:** To assess age-at death distributions, trends in life expectancy and life disparity for Delhi and Kerala from 2010-2018; to assess patterns of life disparity and their relation to rising longevity; to show that, despite having similar life expectancies, life disparity varies between Delhi and Kerala.

**Methods:** Abridged-life table from Sample Registration System from 2010-18 for Kerala and Delhi. Decomposition approach is performed to calculate age-specific contribution to changes in life expectancy and life disparity for each population.

**Results:** Kerala showed inverse relationship between life expectancy and life disparity but not Delhi. Despite having a better Delhi’s life expectancy than Kerala during the study period, Delhi’s life disparity is still higher than Kerala’s. Disparities in life expectancy and life disparity between the two groups are closely related to their differences in age-specific mortality.

**Conclusions:** Given the ongoing demographic transition in India and spatial variations in it, this study is a welcome contribution to our understanding of India’s mortality decline. Our study has revealed that the life disparity in Delhi is higher than that in Kerala. This is because infant mortality in Delhi is higher than in Kerala whereas old age mortality is higher in Kerala than in Delhi.

**Contribution:** Beneficial in allocating healthcare resources to minimize both infants and old deaths and to attain equality in longevity and health.

## 1. Introduction

Life expectancy has been increasing substantially throughout the world over the last few decades. It has been broadly applied as an important indicator in determining the health conditions and mortality profile of a population or a society (Smits & Monden, 2009). Also, it is often used as an early predictor of the societal problems (Hiam et al., 2018). Hence, studies of life expectancy have been important in evaluating the health condition of a population and in implementing effective health policies.

In recent decades, India has been successful in prolonging longevity, but this increase in life expectancy has not been equally spread across the sub-continent, nor even within the different sub-populations. Average mortality levels, usually expressed in terms of life expectancies, are used to track the health of human populations. However, there is significant variation in life expectancy (van Raalte et al., 2018). Many demographers and researchers have suggested that the variations in age at death exhibit health inequality (Tuljapurkar, 2010). Nonetheless, most studies have focused on life expectancy and its association with absolute deprivation than the inequality in the educational, occupational attainment and income (Smits & Monden, 2009). Length of life has been termed the “final inequality”(Tuljapurkar, 2010) but little attention has been paid to the distribution of mortality within a population in Indian settings.

Although life expectancy may still be the best statistic for determining the rate at which the population’s survival improves, life-span variation is a necessary supplement to evaluate equity in survival improvement (van Raalte et al., 2018). However, several measures have been suggested in order to determine lifespan variations or inequalities. For instance, the interquartile range for age at death (Wilmoth & Horiuchi, 1999), the Gini coefficient (V. Shkolnikov et al., 2003), the standard deviation in age at death for ages 10 and older (S10) (Edwards & Tuljapurkar, 2005), and the Theil index of inequality (Smits & Monden, 2009) are all measures of the extent to which deaths are concentrated at one particular age or age group. In the present study, our interest is in life disparity at birth. Life disparity is denoted as *e*^†^ and defined as the average life years lost due to death (Aburto et al., 2020; Vaupel et al., 2011; Zheng et al., 2020). It has often been applied (Vaupel & Canudas-Romo, 2003) to evaluate the variation in age at death that indicates how much lifespans vary among people or individuals. The greater the life disparity, the larger the variation in lifespans and the greater the uncertainty in the age at death. The change (increase or decrease) in life disparity depends entirely on the relation between lives saved “at early ages” and those saved “at late ages” (Vaupel et al., 2011). The shape of the death distribution influences differences in life disparity, with more asymmetric distributions being associated with greater disparities in life (Fernandez & Beltrán-Sánchez, 2022). Another important indicator is the threshold age (*a*^†^), identified by Zhang and Vaupel (2009). Mortality reduction before this age leads to lower disparity, while mortality reduction above this age results in greater disparity (Zhang & Vaupel, 2009). Thus, to comprehend the significant underlying mortality dynamics, it is important to examine the age-specific changes in expectation of life and life disparity.

Earlier studies have found that countries with longer life expectancy also have a more egalitarian lifespan distribution (Vaupel et al., 2011). So, the question arises, does a greater longevity come with the lower disparity in life in India? Over several decades, both men and women in Kerala have enjoyed the longest life expectancy in India. Recently, Delhi, the capital of India with a population of 30.5 million in 2021 (United Nations World Population Prospects, 2021) has overtaken Kerala, and topped the life expectancy ranking with a life expectancy of 75.3 years for both sexes in 2018, compared with 75.3 years for both sexes in Kerala (Office of the Registrar General & Census Commissioner India, 2021). How is life disparity of Delhi compared with Kerala, or in other words, has Delhi attained the longest life expectancy at the loss of life disparity?

Life table data for Delhi were not available till 2009, therefore the present study can only show limited historical trends. Nevertheless, we can see that life expectancy for Delhi has increased from 72 years in 2010-14 to 73.8 years in 2014-18 for males and from 74.7 years in 2010-14 to 77 years in 2014-18 for females (Office of the Registrar General & Census Commissioner India, 2021). Recently, a study conducted by a US research group, showed that life expectancy of Indians could be reduced by nine years due to high level of air pollution (BBC News, 2021). So far, to our knowledge, no study has documented changes in life disparity, together with life expectancy, for Delhi and Kerala, which could enhance our understanding of health conditions in these two states. Life disparity in Delhi has so far not been assessed in any research. Despite that, Delhi and Kerala are high-income societies with the longest life expectancies and the lowest fertility rates in India where TFR is 1.5 in Delhi, TFR is 1.7 in Kerala, compared with a national average of TFR with 2.2 for India as of the latest SRS Report 2018 (Office of the Registrar General & Census Commissioner India, 2018). Nevertheless, they differ in population size, geographical area, urban-rural composition and demographic challenges, as Kerala is more of an aging society. Kerala has stood at the top of life expectancy since the 1970’s, so taking Kerala as a reference while assessing life disparity in Delhi would help to understand the average health situation at a societal level.

Therefore, the aims of the present study are as follow: (1) to assess the death patterns for Delhi and Kerala for the time period 2010-14 and 2014-18, trends in the life expectancy and life disparity for Delhi and Kerala over the time period 2010-2018. (2) to assess the patterns of life disparity and their relation to rising longevity. (3) to show that, despite having similar life expectancies, life disparity varies between Delhi and Kerala.

## 2. Methods

### 2.1 Data source

The sample registration system (SRS) based abridged-life table data has been used in our study from the time-period 2010-2014 to 2014-2018. It was first introduced in 1970, is the primary, continuous and reliable data source that provides mortality and life tables data in India. It is under the support of Office of the Registrar General of India (ORGI), Government of India. Since the life table for Delhi was only available from the year 2010, therefore we have used the life table for both Delhi and Kerala from the time period 2010-2018 in order to conduct our study. The life table data for rural female were not published for the Delhi from 2011-2017, therefore it is not included in our study.

### 2.2 Measures

As we have used the SRS based abridged-life table data for both Kerala and Delhi from 2010-2018. The SRS based abridged-life table data has the open-age category 85+ (85 years and above). So, there is a need to smoothen the mortality data in order to convert into the single age and the open age category to 100+ (100 years and above) for both Kerala and Delhi. Therefore, we have adopted the geom_smooth package in R (version RStudio 2021.09.2) to smoothen the mortality data by fitting it. We have estimated number of deaths in 2010 and 2018 for both Kerala and Delhi by sex and place of residence. And then we have analyzed the trends in life expectancy and life disparity at birth for both Kerala and Delhi by sex and place of residence from the time period 2010-2018. These life expectancy and life disparity measures are comprehensive demographic indicators which are also key indicators for public health policy evaluations (Singh & Ladusingh, 2016) which can also be enhanced by decomposing (Aburto & van Raalte, 2018). Apart from this, the life disparity indicator has showed greater correlation with the other lifespan variation indicators (van Raalte & Caswell, 2013), therefore we have adopted the life disparity measure in our study as an alternative for the other indicators of the lifespan variation (Vaupel et al., 2011; Zhang & Vaupel, 2009; Zheng et al., 2020). We have empirically evaluated the life disparity using the formula given by:

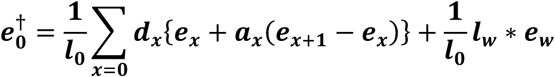

Where the 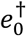 (e-dagger) is denoted as life disparity at age zero or birth; *l*_0_ is the number of survivors at the age zero or ‘0’; *e*_*x*_ is the life expectancy at age *x*; *a*_*x*_ is the average number of the years lived between the age *x* and *x* + 1 by those who died in the interval; *e*_*x*+1_ is the life expectancy at age *x* + 1; *d*_*x*_ is the number of deaths between the age *x* and *x* + 1; *w* is the last age or the limiting age.

We have also established the relationship between life expectancy and life disparity at birth in Kerala and Delhi by sex for rural and urban separately during the time period 2010-2018. This relationship can provide a clear indication of the gaps happening between Kerala and Delhi.

However, the gain in the expectation of life can be attained by saving lives at any age, on the other hand, the loss in life disparity can be obtained by stabilizing the avoidable deaths at both early and late ages. The deaths at early and late ages can be estranged by the threshold ages (a^†^). In order to reduce the disparity in life, there is a need to save the life before the threshold age while on the other hand, the disparity in life could rise when lives are saved after the threshold age (Vaupel et al., 2011). Therefore, we have estimated the trends in the threshold ages (a^†^) which occurred when the following equality holds:

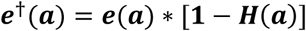

Where ***e***^†^(***a***) is the life disparity at age ***a***; ***e***(***a***) is the life expectancy at age ***a***;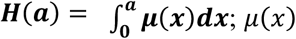 is the hazard, ***H***(***a***) is the cumulative hazard to age *a* where ***H***(**0**) = **0** (Zhang & Vaupel, 2009).

Lastly, we have used the replacement decomposition method proposed by Shkolnikov and Andreev (V. M. Shkolnikov & Andreev, 2010), in order to calculate the age-specific contributions to the changes in the expectation of life and life disparity for both Kerala and Delhi from the time period 2010-2018. The differences in the decomposition approach for the life expectancy and life disparity have been used in our study as:

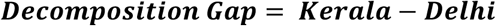

## 3. Results

### 3.1 Age-at death distribution for Kerala and Delhi

Figure 1 (a) demonstrates the number of deaths per 100,000 populations in 2010 and 2018 for Kerala and Delhi by sex for rural. At age zero, the number of deaths is observed greater among Delhi’s males in 2014-18, whereas Kerala’s male mortality was lowest in 2014-18. The death patterns decrease from age zero until before age 25 years, and then it starts increasing and peaked at age 75 years or above and then it came down. The highest number of deaths is seen among Kerala’s males in 2014-18. Figure 1 (b) shows the number of deaths per 100,000 populations in 2010 and 2018 for Kerala and Delhi by sex for urban residence. At age zero, the number of deaths is observed greater among Delhi’s females in 2010-14, whereas Kerala’s male deaths was found to lowest in 2010-14. More people are dying at age 75 years, where Kerala’s male in 2014-18 are at the highest but Delhi’s males have showed the lowest number of deaths at this age. While comparing between figure 1 (a) & (b), we found that higher number of deaths is depicted among rural residence than urban at age zero but at age 75 years, the number of deaths is greater in urban residence compared to rural.

**Figure 1 (a) and (b).**
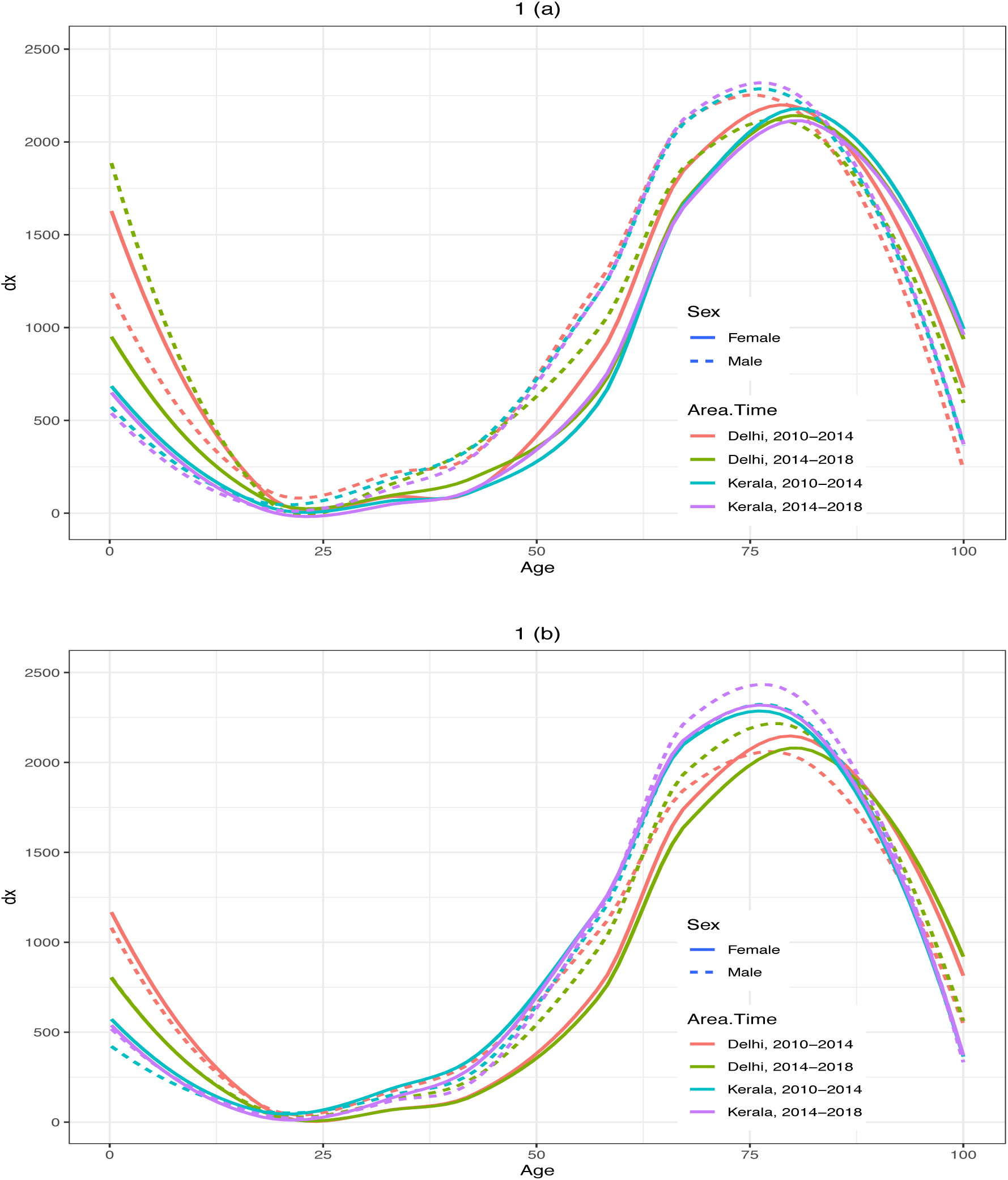
Age-at death distribution (Number of deaths per 100,000 populations) in 2010-14 and 2014-18: Kerala and Delhi for Rural and Urban. **Source:** Authors’ own calculation using the Sample Registration System (SRS) from the Office of the Registrar General of India (ORGI).

### 3.2 Trends in life expectancy and life disparity for Kerala and Delhi

Figure 2 (a) shows the trends in the life expectancy at age 0 for Kerala and Delhi from 2010-2018 by sex and place of residence. The life expectancy at age 0 among males has been showing the increasing trends for Delhi while not much changes has been observed for Kerala. Interestingly, there has been a steadily rise in the female *e*_0_ for Delhi in total residence from 2010-18 but Kerala has been showing small rise in *e*_0_ in 2010-14 and then small drop in *e*_0_ and again marginally increase in *e*_0_ in 2014-18. Despite those larger gaps in the *e*_0_ have been seen between the Kerala and Delhi among both males and females in the rural residence. In urban residence, the *e*_0_ is showing the increasing trends among males and females for Delhi where Delhi has overtaken Kerala among males in 2011-2015 but not for urban female where the *e*_0_ for Kerala is still higher. Although the gaps in *e*_0_ between Kerala and Delhi is reducing for urban female, which might be overtaken in future.

**Figure 2 (a) & (b):**
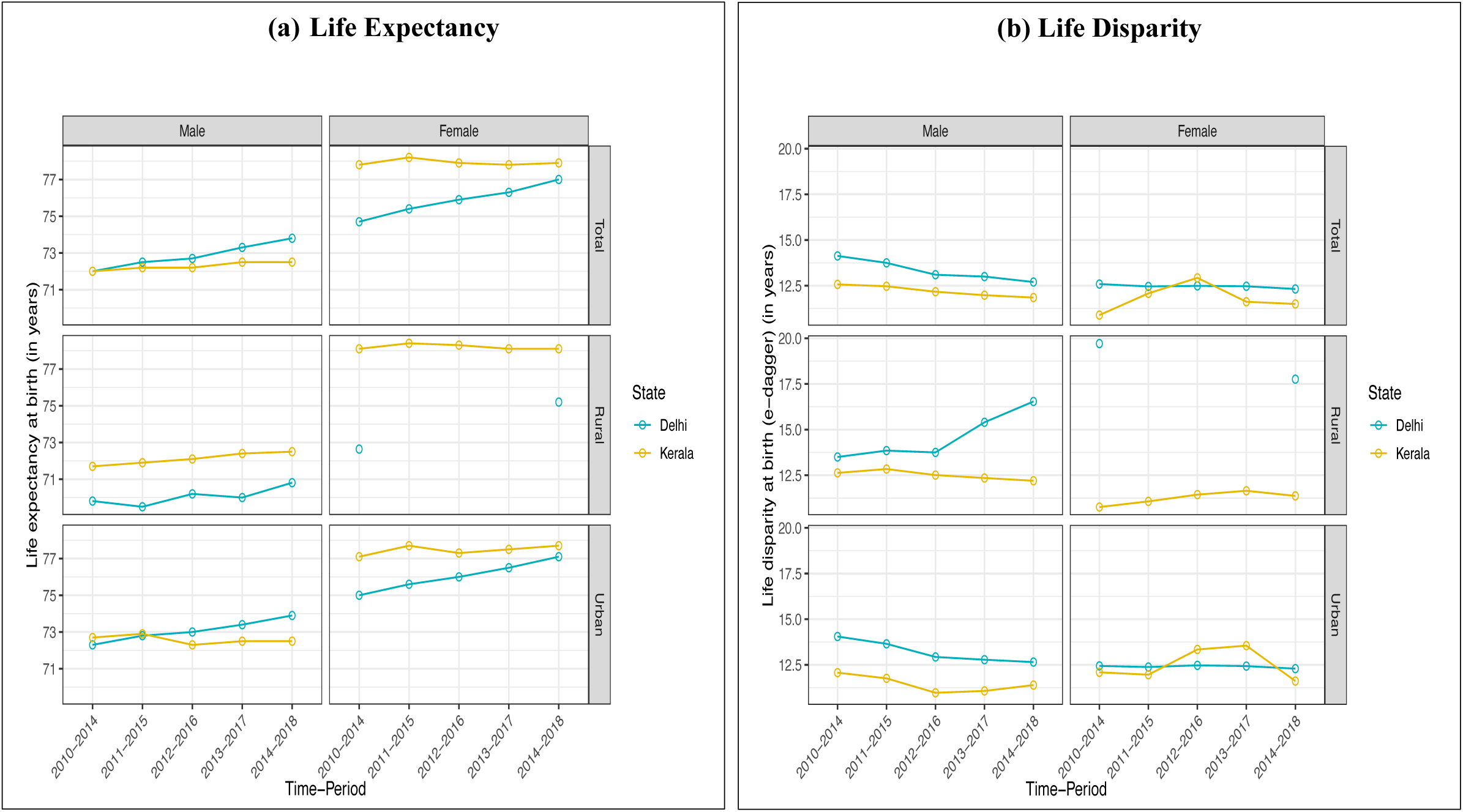
Trends in life expectancy (LEB) and life disparity at birth (LDB) in Kerala and Delhi during the period 2010-2018. **Note:** Rural female data for Delhi were not published during 2011-2017. **Source:** Authors’ own calculation using the Sample Registration System (SRS) from the Office of the Registrar General of India (ORGI). **Abbreviations:** LEB-Life expectancy at birth; LDB-Life disparity at birth.

However, Figure 2 (b) presents the trends in life disparity at birth for Kerala and Delhi from 2010-2018 by sex and place of residence. The result clearly showing that the life disparity for Delhi has been higher than Kerala except for total female in 2012-16 and for urban female from 2012-17. The study reveals that the male has higher life disparity than female in total residence for Delhi while in Kerala the female life disparity was initially lower than male, but it rises and then it has declined. In case of rural residence, Kerala showing higher male life disparity than female throughout the period. Urban male life disparity for Delhi is showing declining trend while there is not much change is observed for female, but still the urban male life disparity is higher than female. Kerala’s female life disparity in urban residence has increased from 2011-17 and then it has declined in 2014-18. Strikingly, urban female disparity for Kerala is higher than male.

### 3.3 Relationship between life expectancy and life disparity for Kerala and Delhi

Figure 3 shows the relationship between the life expectancy at birth and life disparity at birth for Kerala and Delhi for 2010-14 and 2014-18. It is interesting to see that the life disparity at birth goes down as the life expectancy at birth increases for Kerala among both male and female, but all indicators for Delhi are lying all above the regression line except for rural male in 2010-14 which is lying just below the regression line. Rural females of Kerala showing lowest life disparity with increase in life expectancy.

**Figure 3:**
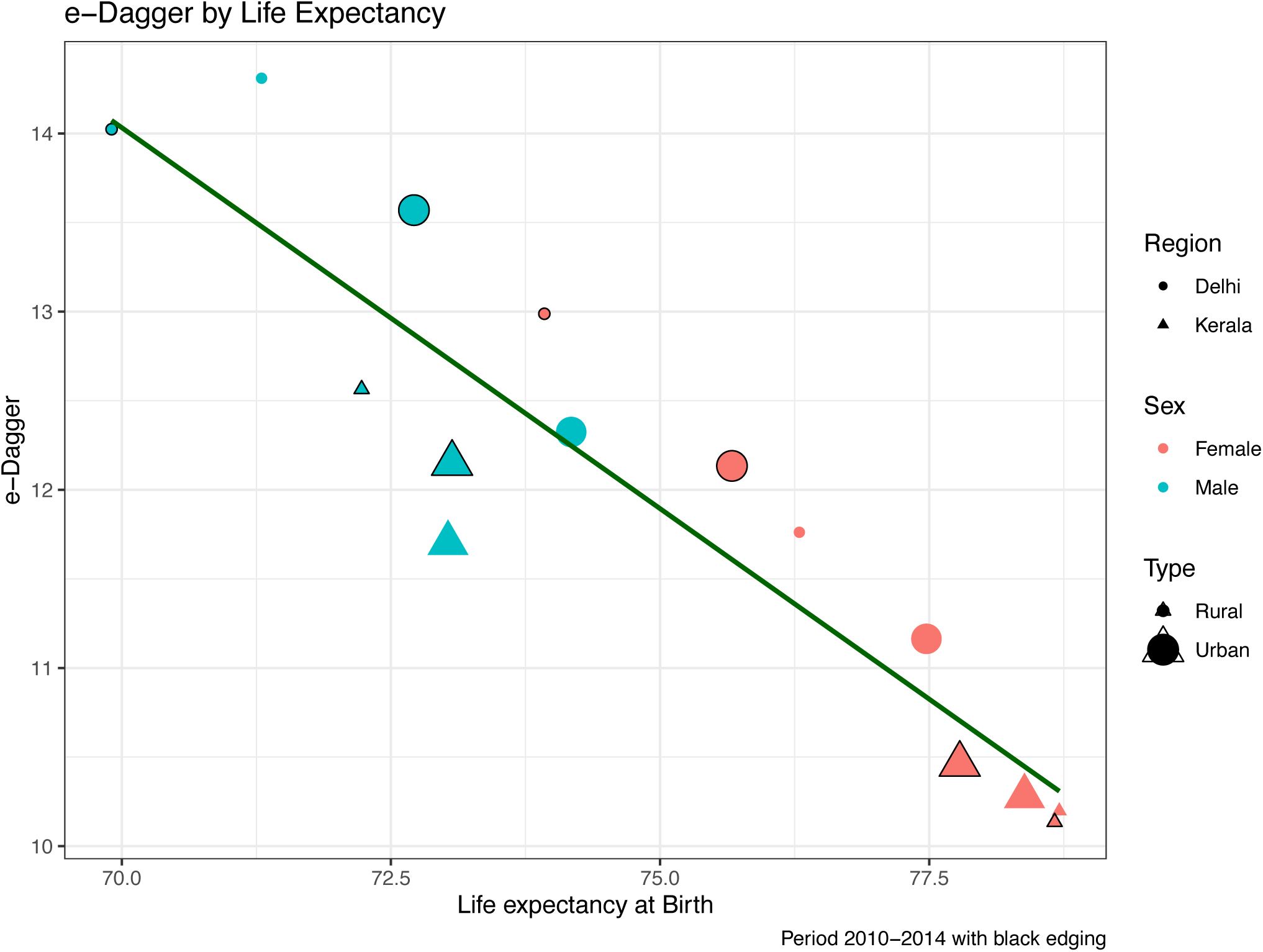
The relationship between life expectancy and life disparity at birth in Kerala and Delhi during the time period 2010-2018. Note: Rural female data for Delhi were not published during 2011-2017. **Source:** Authors’ own calculation using the Sample Registration System (SRS) from the Office of the Registrar General of India (ORGI). **Abbreviations:** LEB-Life expectancy at birth; LDB-Life disparity at birth.

### 3.4 Trends in threshold age for Kerala and Delhi

Figure 4 indicates the trends in threshold age for Kerala and Delhi from 2010-2018 by sex and place of residence. For most of the observed years, Kerala has been showing the higher threshold ages than Delhi except for males in total and urban residences. The highest threshold age can be seen among female of Kerala belonging to rural residence in 2013-17 whereas Delhi’s male showing the lowest threshold age belonging to rural residence in 2011-15. In 2014-2018, Kerala’s male threshold age was 72.39 years, implying that lowering death before 72.39 years would result in a decrease in life disparity, whilst reducing mortality after 72.39 years would result in an increase in life disparity.

**Figure 4:**
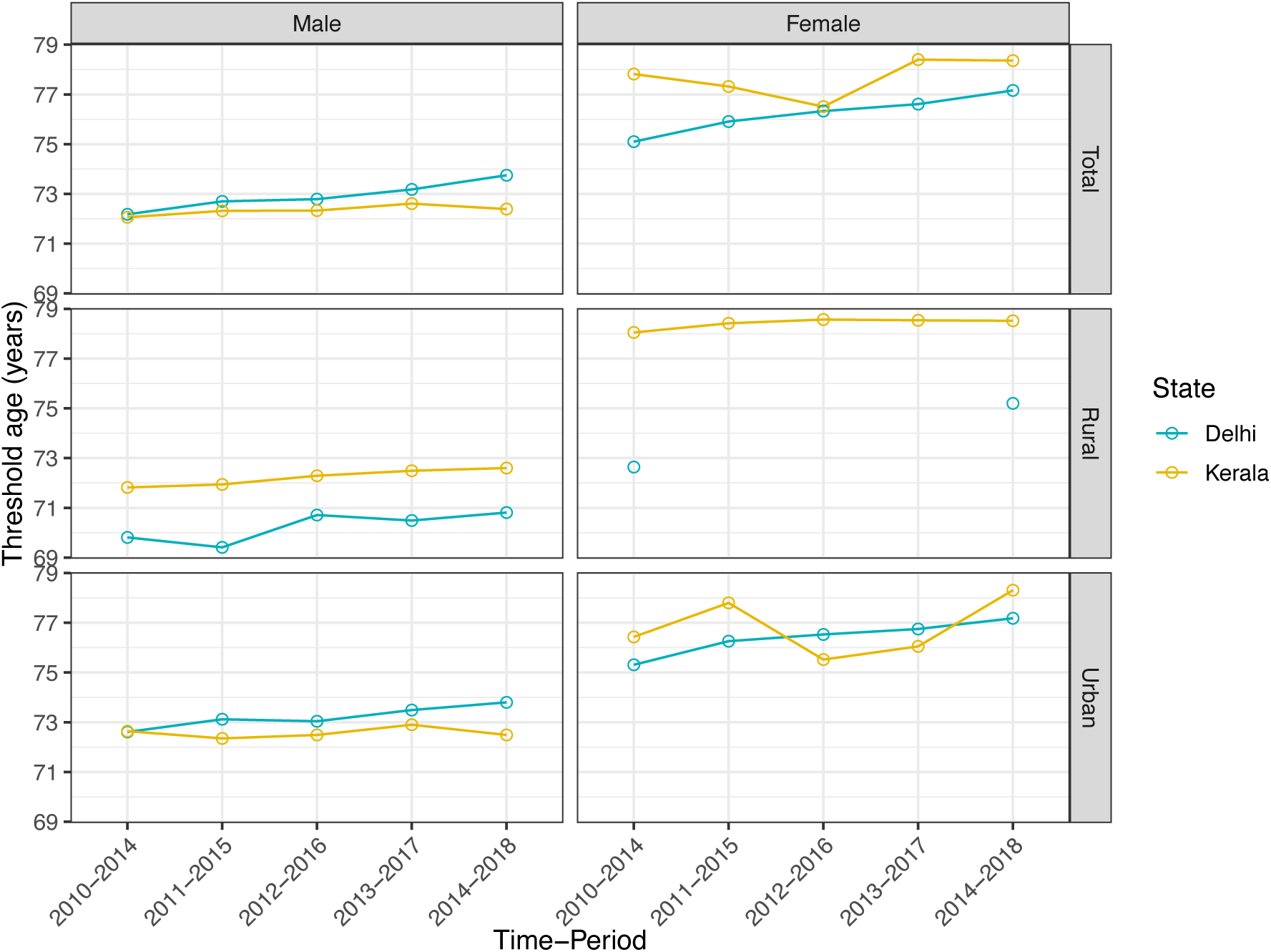
Trends in threshold ages (a^†^) in Kerala and Delhi during the period 2010-2018. Note: Rural female data for Delhi were not published during 2011-2017. **Source:** Authors’ own calculation using the Sample Registration System (SRS) from the Office of the Registrar General of India (ORGI).

### 3.5 Decomposition results of the life expectancy between Kerala and Delhi

Figure 5 depicts the age-specific decomposition of the differences in life expectancy between Kerala and Delhi by sex and residence. The gaps in life expectancy are indicated by the red lines (i.e., Kerala minus Delhi, abbreviated as KRL-DEL). The result clearly shows that the gaps in life expectancy among males are lower than females in total and urban residence. Delhi’s advantage in male life expectancy in 2010-14 was mainly due to the lower death rates in the age group 25-59. Since 2010-14, Delhi has overtaken Kerala in male life expectancy in the total residence; this is mainly because Delhi has caught up with Kerala in mortality and outperformed Kerala in reducing the mortality at the 60-84 age groups. For most years, males for all ages belonging to total residence are negatively contributing to the KRL-DEL gap. This means that consistently, Kerala had higher male mortality in that respective age group.

**Figure 5:**
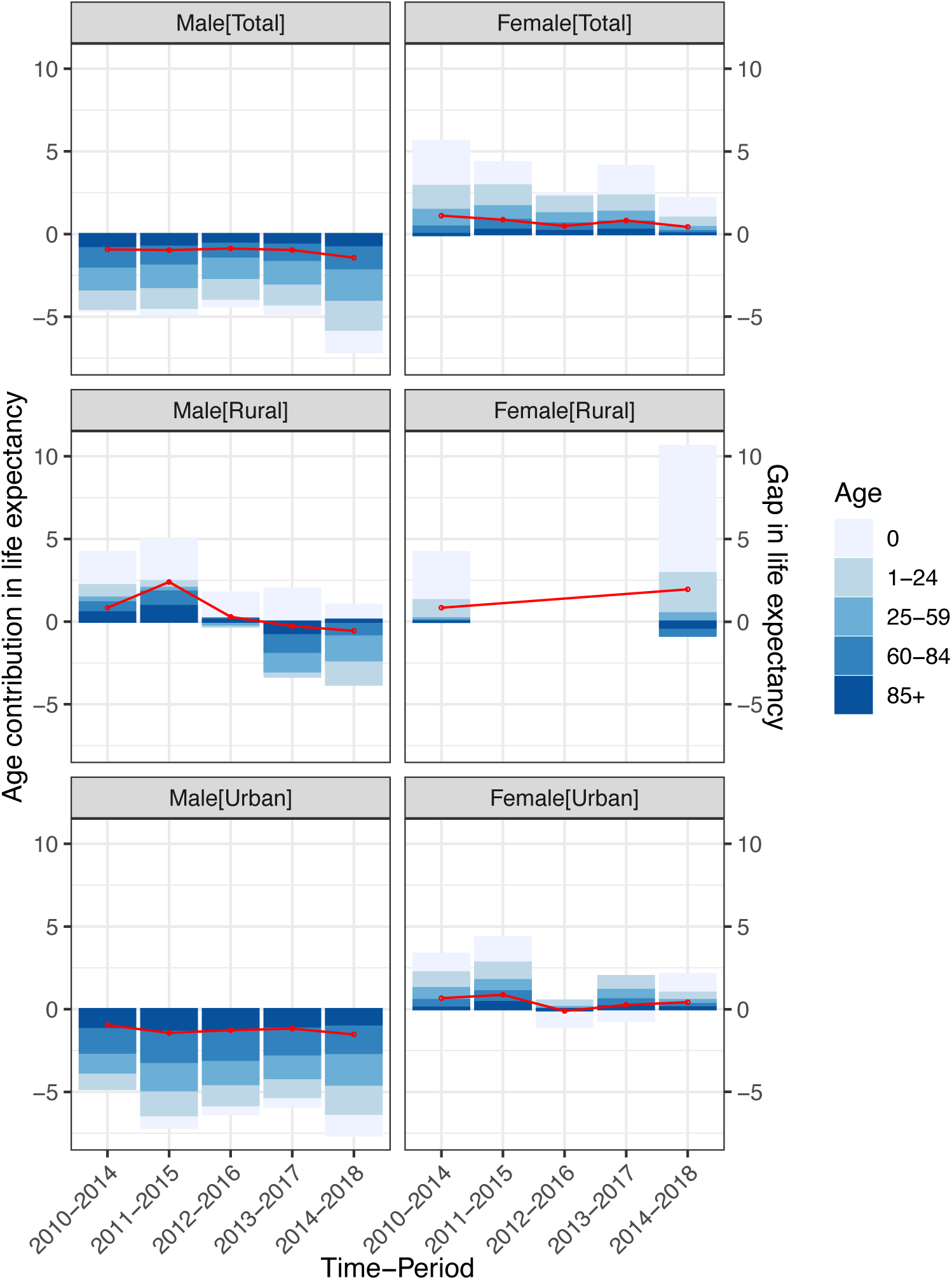
Decomposition results showing the gaps in life expectancy between Kerala and Delhi during 2010-2018. **Notes:** The red lines are showing the gaps in the life expectancy between Kerala and Delhi (Kerala minus Delhi); Rural female data for Delhi were not published during 2011-2017. **Source:** Authors’ own calculation using the Sample Registration System (SRS) from the Office of the Registrar General of India (ORGI).

However, the trend in life expectancy gaps is depicted to be slightly decreased among females while increasing trend is seen among rural females but there is not much change is observed among urban females. In the most of the years, Kerala’s female had longer life expectancy than Delhi, this was because Kerala has lower female mortality. As Delhi’s female mortality has not much improved to the level of Kerala, the KRL-DEL gap has not showed much change. Currently, Delhi’s disadvantage in female mortality which has offset Kerala’s advantage in female mortality at the all ages, as a result the gap is very wide. It is also worth noting that Delhi’s remarkable decrease in mortality rates among those aged 60-84 had a substantial impact on both males and females.

### 3.6 Decomposition results of the life disparity between Kerala and Delhi

Figure 6 visualizes the age-specific decomposition of the differences in life disparity between Kerala and Delhi by sex and place of residence. Unlike increasing longevity can be attained by saving lives of people at all ages, the change in life disparity is driven by the balance between changes in mortality at young and old age groups.

**Figure 6:**
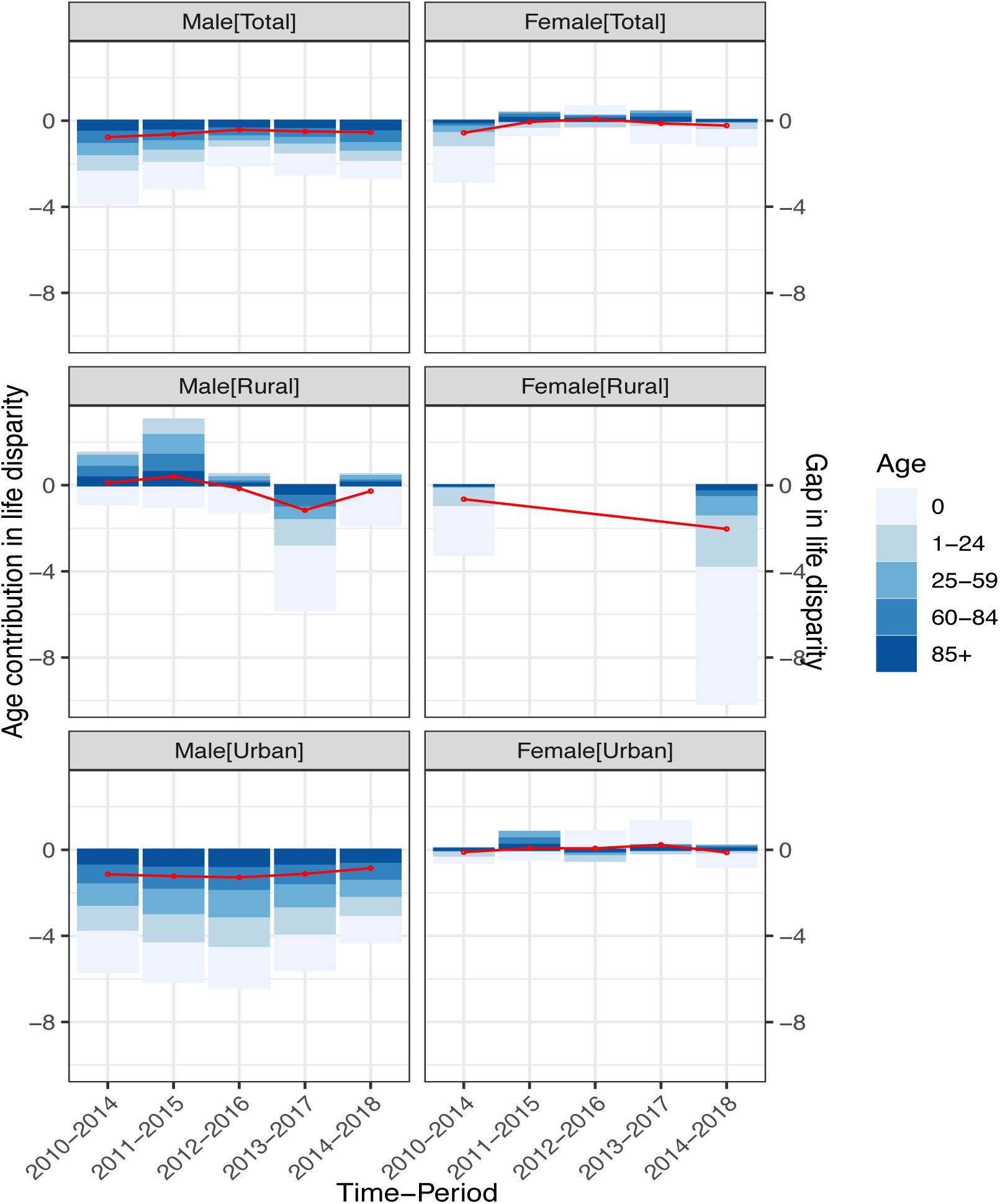
Decomposition results showing the gaps in life disparity between Kerala and Delhi during 2010-2018. **Notes:** The red lines are showing the gaps in the life disparity between Kerala and Delhi (Kerala minus Delhi); Rural female data for Delhi were not published during 2011-2017. **Source:** Authors’ own calculation using the Sample Registration System (SRS) from the Office of the Registrar General of India (ORGI).

As shown, Delhi has higher life disparities than Kerala for both males and females except for some age groups. This was mainly contributed by Delhi’s greater mortality rates at age zero, 1-24 and 25-59 age groups. In other words, higher male and female premature mortality rates in Delhi contributed to its higher life disparities. Over the time, Delhi has achieved great improvements in reducing premature mortality, which is resulting in a significant fall in life disparity. The gap in the life disparity between Kerala and Delhi has narrowed down among rural female but not much change is observed among others.

## 4. Discussion

Our study takes into account not just a life expectancy, as it is the most fundamental statistic applied to represent health and mortality status, as well as another dimension of mortality by assessing life disparity, providing a more comprehensive understanding of health conditions in Delhi. Likewise, comparisons have been made with trends in Kerala by sex and place of residence, that has set a record for life expectancy since the 1970s (Thomas & James, 2014) and has furthermore achieved remarkable success in lowering life disparity. This comparative study with Kerala has improved our comprehension of Delhi’s health predicament in a national and subnational setting.

Between 2010-14 and 2014-18, life expectancy and life disparity in Kerala had a typically negative correlation in a long-term trend, indicating that Kerala may reach higher life expectancies while also having lower life disparities. While Delhi showed a positive correlation, which means that the life disparity has also increased even with an increase in life expectancy. The current findings for Kerala that showed a strong negative association between the life expectancy and life disparity. However, the life expectancy has historically been substantially inversely associated with lifespan variation (Colchero et al., 2016; van Raalte et al., 2018; Vaupel et al., 2011), this is no longer the case for Delhi.

Furthermore, the study’s decomposition analyses assessed the age-specific contributions to the increase in life expectancy and the change in life disparity in the two different populations. The improvement in life expectancy at this stage of development in both high-income societies, Delhi and Kerala, was mostly due to lower mortality rates in the adult and elderly age groups, with minor contributions from new-borns, children, and teenagers. The drop in life disparity for Kerala was due to a net effect of lower premature mortality, which negated the effect of lower mortality among the elderly people.

However, the contributions of different ages to the overall differences in life expectancy and life disparity between Delhi and Kerala were studied in this study. The findings demonstrate that, despite having a better Delhi’s life expectancy than Kerala during the study period, Delhi’s life disparity is still higher than Kerala’s. The disparities in life expectancy and life disparity between the two groups are closely related to their differences in age-specific mortality, according to this comparison. Delhi has made significant progress in lowering premature mortality for men, narrowing the life expectancy and disparities gap with Kerala during the given period.

Notwithstanding, our study has highlighted three significant findings. First, in recent years, Delhi’s life expectancy advantages for both males and females have been driven by a remarkable drop in mortality among the older adult population (85+ years), which, on the other hand, has widened the life disparity gap with Kerala. Second, the recent disappearance of the male life disparity was due to Delhi’s reduced mortality rates across practically all age categories, not because the two cultures had identical male mortality rates. other hand, has widened the life disparity gap with Kerala. Delhi’s lower premature mortality has narrowed the gap compared to Kerala, while its lower mortality after the threshold age has widened it. Third, the widening of the KRL-DEL female life disparity can be attributed to Delhi’s reduction in mortality among the older adult populations, which has more than offset the improvement in premature mortality. A different narrative is presented between United States and United Kingdom comparison (V. M. Shkolnikov et al., 2011) than our comparative results between KRL-DEL. Conversely, our study has found that Delhi has a similar life expectancy to Kerala, but a larger life disparity due to Delhi’s reduced mortality after 65. These two variants of the life expectancy-disparity nexus emphasize the importance of looking at age-specific determinants in order to understand the underlying mortality dynamics.

Our study has again confirmed with the previous studies (Seaman et al., 2016; Smits & Monden, 2009; van Raalte et al., 2018) that even in societies with equal levels of life expectancy, differing levels of life disparity can be identified. A study by (Smits & Monden, 2009) concluded that countries that attained a specific level of life expectancy earlier in time were more likely to experience higher levels of inequality, based on life tables for 212 countries. Nonetheless, the outcomes of a study conducted by (Seaman et al., 2016) are contradictory: Because of its lower mortality among older adult populations but greater premature mortality among adults, Scotland, which had caught up to England and Wales in terms of life expectancy, had bigger variations in lifespan. Likewise, the KRL-DEL comparison reveals that Delhi has attained a similar level of life expectancy as Kerala later on time, but with a larger life disparity, owing to Delhi’s lower old-age mortality.

Our findings on the growing significance of mortality among older adults in determining life disparity in Delhi suggest that the positive link between life expectancy and life disparity may shift to a negative one, with both increasing simultaneously. If old age mortality is even further delayed or lowered, while mortality rates at younger ages and middle-age adults may stagnate or begin to climb (van Raalte et al., 2018), such an inversion is possible. Earlier studies in Denmark (Brønnum-Hansen, 2017) and Finland (van Raalte et al., 2014) have suggested that death rates at younger ages may be rising or stagnating which can be associated to various socioeconomic conditions, which may contribute to disparities in lifespan variation, — particularly among certain subpopulations. As a result, it is important to introduce policies for justifying social arrangements. Furthermore, another study (Seligman et al., 2016) has identified that the causes of death that resulted in larger life disparity were distinct from the causes of death that resulted in higher life expectancy. This would be helpful in promoting health at a younger age which could increase equality while controlling cardiovascular and cancer diseases would play a key role in increasing longevity. Hence, it is essential to allocate health care resources in order to limit the cause of mortality at both young and older adults, allowing the society to attain both longevity and equality.

## 5. Limitations

There are certain limitations in our study. First, the lifetable data for Delhi is only available from 2010-14, therefore, our study could not analyze the historical trends in life expectancy and life disparity, but that would not impact our results. Though the age-specific death rate (ASDR) data from the SRS Reports were available for Delhi from 2004 to 2019, we could not use it. This is because some of the ASDR estimates for specific age-groups for Delhi for almost every year are ‘zero’. Therefore, we did not perform using this data; however, it could better understand long-term trends. Second, the data is not available for Rural Delhi from 2011-15 to 2013-17 because Delhi is predominantly urban. In the SRS data, there are only 10 units from Rural Delhi, with such a small sample, the sampling errors become larger and the estimates become erratic. Therefore, it is not prudent to obtain estimates for Rural Delhi separately. Third, due to data limitation, only age, sex and place of residence were included in the analysis, without looking into the causes of death. However, the decomposition results can be studied to evaluate the contributions of different diseases to changes in life expectancy and life disparity if more micro death registration information is available. The study by age and cause could provide more information for future public health strategies. Fourth, our study has more focused on the life disparity and life expectancy rather than looking at patterns within different socioeconomic factors. Thus, future studies could make an effort to mitigate this issue, which would be extremely beneficial. Lastly, the impact of migration cannot be assessed due to unavailability of information on migrants.

## 6. Conclusions

Given the ongoing demographic transition in India and spatial variations in it, this study is a welcome contribution to our understanding of India’s mortality decline. Our study has revealed that the life disparity in Delhi is higher than that in Kerala. This is because infant mortality in Delhi is higher than in Kerala whereas old age mortality is higher in Kerala than in Delhi. Since the distribution of deaths is bimodal, the modes being age ‘0’ and a high age (at an old age), as infant and early childhood mortality falls, life disparity declines. With the number of older people in Delhi and Kerala increasing, greater attention should be devoted to their medical treatment and healthcare demands. Further efforts should be undertaken to improve premature mortality in Delhi in order to reduce the life disparity. Preventing premature mortality requires targeted and effective health policy measures. Individuals’ awareness of their less-uncertain lifetimes and life plans, as well as consequences on public policies, are influenced by reductions in life disparity and rises in life expectancy. With the increased likelihood of living a longer life, it is advised that retirement ages be adjusted to keep up with the increase in life expectancy. Also, the definitions of “aged” and “dependent” may need to be reconsidered, particularly in terms of “prospective” ageing metrics. Extension of the retirement age may be a strategy of generating human and social capital in the future for Delhi and Kerala, which have the highest life expectancies and lowest fertility rates in India.

## Data Availability

The study utilises secondary source of data which is freely available in public domain through
https://censusindia.gov.in/vital_statistics/Appendix_SRS_Based_Life_Table.html

https://censusindia.gov.in/vital_statistics/Appendix_SRS_Based_Life_Table.html

## 7. Acknowledgments

During the final stage of this work, I gratefully acknowledge Prof. P.M. Kulkarni (Former Professor at CSRD, JNU, New Delhi, India) for his insightful comments and recommendations. I gladly applaud Dr. José Manuel Aburto for his significant remarks and recommendations. Finally, I would like to express my gratitude to Prof. Nandita Saikia (my PhD advisor) for her encouragement and inspiration in the field of mortality research.

